# Analysis: The potential health and economic impact of dexamethasone treatment for patients with COVID-19

**DOI:** 10.1101/2020.07.29.20164269

**Authors:** Ricardo Águas, Adam Mahdi, Rima Shretta, Peter Horby, Martin Landray, Lisa White, the CoMo Consortium

## Abstract

Dexamethasone has been shown to reduce mortality in hospitalised COVID-19 patients needing oxygen and ventilation by 18% and 36%, respectively. Here, we estimate the potential number of lives saved and life years gained if this treatment would be rolled out in the UK and globally, as well as its cost-effectiveness of implementing this intervention. We estimate that, for the UK, approximately 12,000 [4,250 - 27,000] lives could be saved by January 2021. Assuming that dexamethasone has a similar effect size in settings where access to oxygen therapies is limited, this would translate into approximately 650,000 [240,000 - 1,400,000] lives saved globally. If dexamethasone acts differently in these settings, the impact could be less than half of this value. To estimate the full potential of dexamethasone in the global fight against COVID-19, it is essential to perform clinical research in settings with limited access to oxygen and/or ventilators, e.g. in low and middle-income countries.

---

Coronavirus disease 2019 (COVID-19), emerged in late 2019 and is either asymptomatic or causes only mild symptoms in most individuals^2^. However, a significant number of individuals, especially amongst the elderly, develop a more severe form of the disease and require hospital care. A further subset of these patients requires oxygen therapy or ventilatory assistance, creating a high demand for long-term hospital care in intensive care units (ICUs). In the UK, the case fatality rate among those admitted to hospital with COVID-19 is more than 26%; this rises to more than 37% in patients who require mechanical ventilation^3^. An extensive search for potential drugs with which to treat COVID-19 is underway. Dexamethasone has emerged as a standout therapeutic candidate reducing mortality in hospitalised COVID-19 patients needing oxygen and ventilation by 18% and 36%, respectively^1^. Thus, the adoption of a dexamethasone treatment protocol for patients requiring respiratory support could potentially lead to a significant number of lives saved over the course of the next 6 months of the pandemic.

Lockdown measures are being eased around the world, creating concern about potential resurgences in COVID-19^4^ activity. As stringent transmission-reducing interventions are lifted, the expectation is that a second epidemic peak becomes inevitable, and with it a significant increase in mortality. Dexamethasone is an affordable medicine available as a generic product that has been routinely used in hospital settings globally and is thus, assuming global availability is maintained^5^, perfectly placed as a candidate standard therapeutic option for COVID-19 patients with respiratory distress, which could significantly reduce the future mortality burden of this disease in a cost-effective way.

## COVID-19 exposure and mortality in the UK

Estimates of the future burden of COVID-19, and therefore the impact of any treatments for COVID-19, are highly dependent both on viral and human behavioural factors. Both are riddled with uncertainty and are changing as the pandemic evolves. Several biological parameters describing how the virus transmits between individuals and how infectivity progresses following infection remain elusive. A critical metric that reflects both how transmissible and lethal the virus can be is the infection fatality ratio (IFR). Until very recently, obtaining a clear picture of how many people have been infected was only feasible in small, well contained outbreaks, such as those on cruise ships^6^. Even in those instances, the narrow time window for good Polymerase Chain Reaction (PCR) testing sensitivity meant even the best estimates for IFR were changing on a weekly basis. Establishing the relationship between deaths and infections is crucial to enable reliable mortality burden predictions for the next wave of the epidemic.

Since May 2020, serological studies have been conducted, in several countries, to measure the proportion of the population that has been exposed to the virus, including in the UK^7^. We now know that at least 7% of the UK population has been exposed to COVID-19, while around 40,000 patients have died from the disease^8^. Of these patients, if 59% had received oxygen and 17% had been ventilated^1^ then, using the reported reduction in mortality risk ^1^, approximately 6,700 individuals would have survived had they been given dexamethasone, according to this simple calculation: [(0.18 × 0.59) + (0.36 ×0.17)] × 40,000 = 6,696.

Given a reasonable IFR estimate, we can project different SARS-COV-2 transmission progression scenarios for the next 6 months, along with their respective mortality burdens, notwithstanding a great deal of uncertainty around what future social distancing measures will be taken and how the population will adhere to those measures. Here, we describe a simple and transparent approach to determine the potential benefits of implementing a standard COVID-19 treatment protocol with dexamethasone in terms of lives saved, life years gained, and cost per life saved. In simple terms, given a country’s population age structure, we use age-dependent estimates for the relationships between infection and mortality and infection and hospitalisation to extrapolate the expected number of hospital admissions and COVID-19 deaths given a specific projected number of infections over the next 6 months. Assumptions about oxygen treatment requirements and expected probabilities of mortality given the use of dexamethasone are clearly stated and can be altered to explore alternative options for these values.

## Results

### Potential lives saved with dexamethasone treatment

We consider the treatment of patients hospitalised with COVID-19 disease, as defined in^3^ as when a patient presents with (i) typical symptoms (e.g. fatigue with fever and muscle pain, or respiratory illness with cough and shortness of breath); and (ii) compatible chest X-ray findings (consolidation or ground-glass shadowing); and (iii) alternative causes have been considered unlikely or excluded (e.g. heart failure, influenza). However, the diagnosis remains mainly based on clinical symptoms and is ultimately at the managing doctor’s discretion.

Working from the population age distribution in the UK as reported by the UN 2019 Revision of World Population Prospects^9^, we assume that among those who are infected, the proportion hospitalised is as estimated for the French population^10^. We also assume that the hospitalised case fatality ratio by age is correlated with that estimated for the French population^10^. More specifically, we developed a hospitalised patient treatment pathway similar to the one in^11^, which can be distilled into a decision tree algorithm (Figure 1) driven by the parameters listed in Table 1 and Supplementary Table 1.

**Table 1:** Definitions of the values used for the calculation and, where relevant, the ranges of values used for the sensitivity analysis.

| Definition | Symbol | Value | Range |  | Source |
| --- | --- | --- | --- | --- | --- |
|  |  |  | Lower | Upper |  |
| Proportion of population expected to be exposed during the 6-month period | $\alpha$ | 0.07 | 0.05 | 0.15 | <sup>8</sup> |
| Level of access to appropriate respiratory support (as a proportion) | $\kappa$ | 1 | 0.5 | 1 | - |
| Increase in fatality rate if no access to appropriate respiratory support | $\psi$ | 1 | 1 | 2 | - |
| Probability of hospitalised patient needing oxygen (either non-invasive or ventilator) | $p_{O_2}$ | 0.76 | 0.2 | 0.9 | <sup>1</sup> |
| Probability of an hospitalised patient who receives oxygen needing a ventilator if on dexamethasone | $p_{vd}$ | 0.22*<br>0.87 | 0.04 | 0.5 | <sup>1</sup> |
| Probability of an infected patient who receives oxygen needing a ventilator if not on dexamethasone | $p_v$ | 0.22 | 0.05 | 0.6 | <sup>1</sup> |
| Maximum death probability for a hospitalised infection | $\delta_H$ | 0.3 | - | - | <sup>3</sup> |
| Maximum death probability for a hospitalised infection requiring non-invasive oxygen | $\delta_{O_2}$ | 0.4 | - | - | <sup>3</sup> |
| Maximum death probability for a hospitalised infection requiring a ventilator | $\delta_v$ | 0.66 | - | - | <sup>3</sup> |
| Reduction in mortality for patients requiring non-invasive oxygen if on dexamethasone | $\epsilon_{O_2}$ | 0.2 | - | - | <sup>1</sup> |
| Reduction in mortality for patients requiring a ventilator if on dexamethasone | $\epsilon_v$ | 0.36 | - | - | <sup>1</sup> |
| Incremental number of days spent in a general ward bed for a survivor that required non-invasive oxygen | $ids_{O_2}$ | 4.5 | 2 | 7 | <sup>15</sup> |
| Incremental number of days spent in a general ward bed for a survivor that required ventilation | $ids_v$ | 5 | 3 | 8 | <sup>15</sup> |
| Incremental number of days spent in an ICU bed for a survivor that required ventilation | $idc_v$ | -1 | -2 | 3 | <sup>15</sup> |

**Figure 1:**
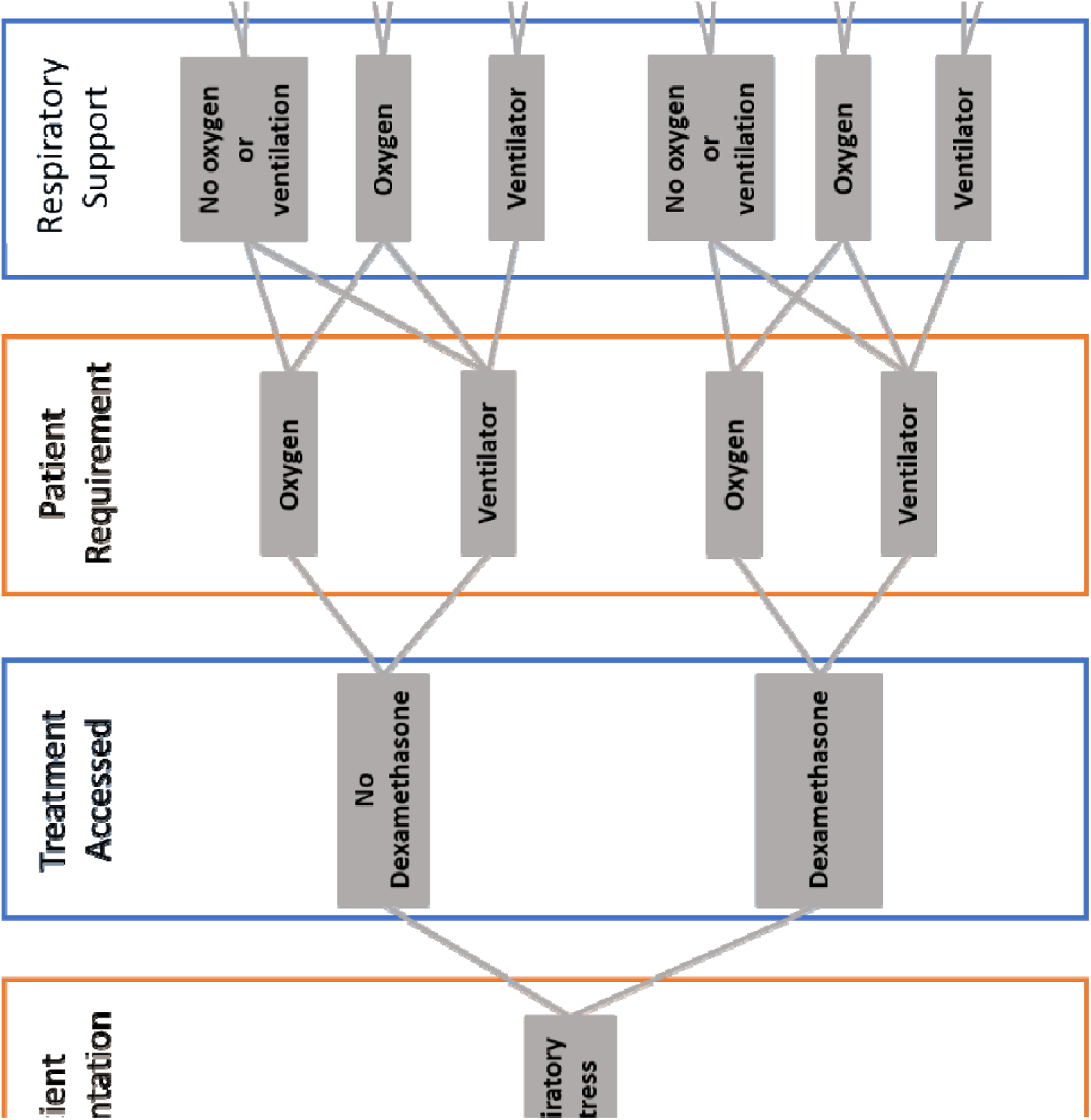
Patient pathway diagram for the calculation of COVID-19 mortality for a given exposure level.

**Figure 2:**
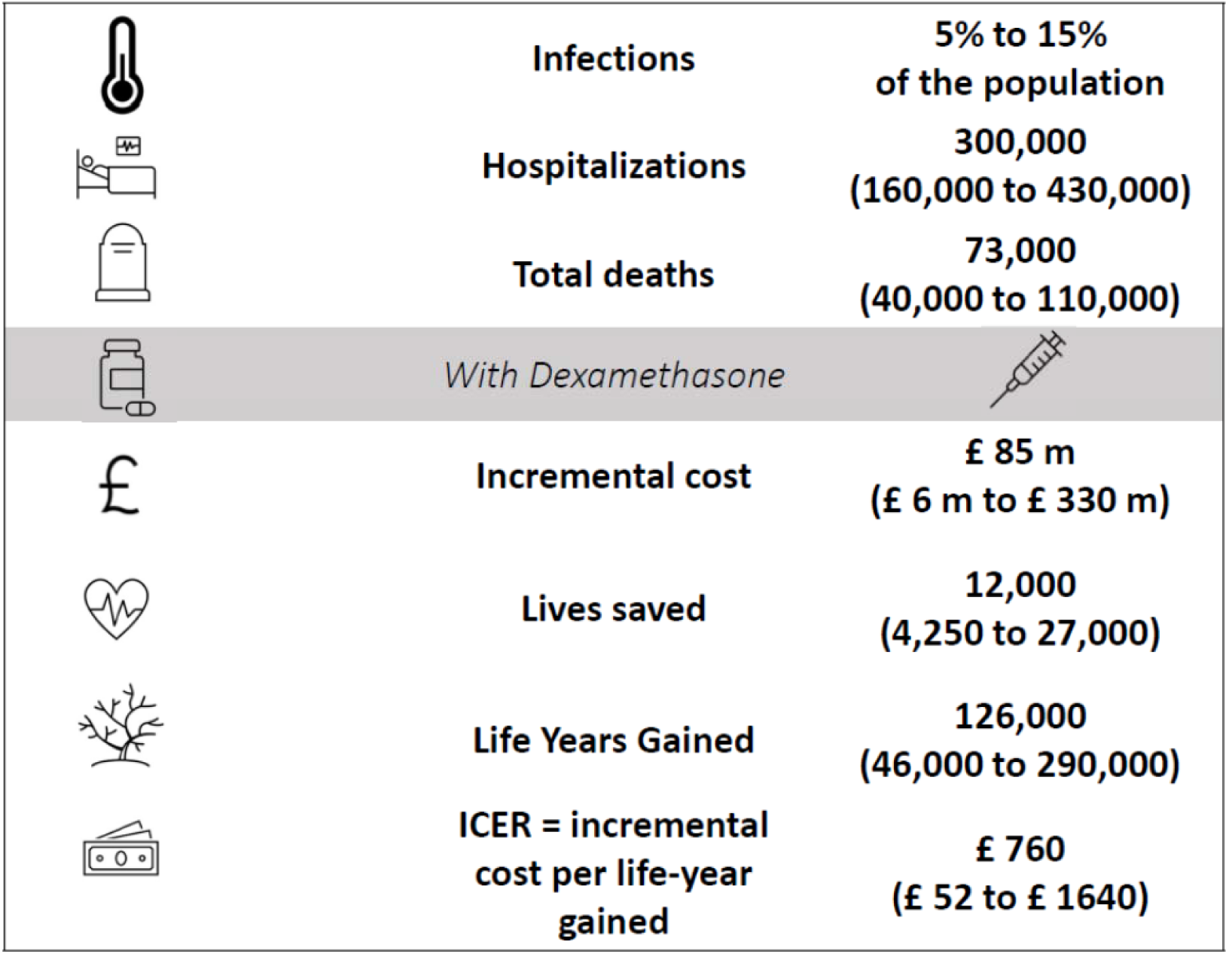
Expected impact of dexamethasone in the UK over the next six months for the range of scenarios explored. The value and range quoted for each outcome represent the median and 5^th^ and 95^th^ percentiles of the sensitivity analysis outcomes, i.e. 90% double-sided confidence intervals.

This algorithm can straightforwardly translate a projected number of total infections into an expected number of hospitalisations by age group. For a range of likely exposure levels over the next 6 months, we can estimate the respective expected mortality, as was done in^12^, by applying the probabilities in Table 1 and following the branching processes in Figure 1. We make this estimate under two scenarios: (1) patients do not receive dexamethasone; (2) patients receive dexamethasone if they meet the criteria for the treatment as described in^1^. The number of potential lives saved by adopting dexamethasone as a standard COVID-19 treatment is simply the difference between these figures. We estimate that, for the UK, approximately 12,000 (4,250 - 27,000 90% confidence interval) lives could be saved over the next 6 months (July-December 2020), under perfect access to treatment.

However, whilst in the UK it is reasonable to assume patients will have perfect access to whatever treatment they require, including dexamethasone, that assumption is unrealistic for many low and middle-income countries (LMICs). Of note, in scenarios where access to oxygen therapy or ventilatory support is limited, the effect of dexamethasone on patient outcome is unknown. It is therefore essential to perform clinical research in LMICs, where access to oxygen and ventilators is limited, to estimate the full potential of dexamethasone in the global fight against COVID-19.

To address this uncertainty, we simulated the use of dexamethasone if a patient meets the oxygen prescribing target described in^13^, whether or not oxygen is available. Whilst the impact of dexamethasone on patients who require but cannot access supplemental oxygen or invasive mechanical ventilation is not known, it is reasonable to assume that some mortality benefit would arise from the reduction of inflammation and consequent decrease in the probability of progressing to respiratory failure. We estimated the potential number of lives saved under two assumptions: (1) dexamethasone has the same relative impact on mortality in patients who do not receive the respiratory support they require as in those who do; (2) dexamethasone has no impact on patient outcome if the patient does not receive the oxygen or ventilation they need. When including access to respiratory support as a covariate in the sensitivity analysis, we obtain an estimated approximately 650,000 (240,000 - 1,400,000 90% confidence interval) potential lives saved under the first assumption and 390,000 (130,000 - 1,000,000 90% confidence interval) lives saved under the second assumption.

Dexamethasone is an affordable drug that has been on the market for many years^14^. Its use can substantially alter outcomes for patients with COVID-19 and by doing so will impact hospital occupancy management and overall cost of treatment. Here, we also explore the economic ramifications of adopting dexamethasone as a default option for treating patients with COVID-19 and provide metrics for the cost-effectiveness of this conceivable health policy change in the UK. To accomplish this, we procured daily hospital patient costs per treatment given as well as data on length of hospital stay for patients with different hospital pathways and disease outcomes – see Methods for more details. According to National Institute for Health and Care Excellence (NICE) directives, interventions with an ICER of less than £20,000 per QALY or life year gained are considered to be cost effective^18^.

In the UK, we estimate a total incremental cost of £85,000,000 (£6,000,000 - £330,000,000 90% confidence interval) over the next 6 months, which equates to £8,200 (£650 - £17,500 90% confidence interval) per life saved and £760 (£52 - £1640 90% confidence interval) per life year gained, making dexamethasone treatment a clearly cost-effective option (**Error! Reference source not found**.). We have refrained from presenting these estimates for the rest of the world due to the large variance in hospital patient management, hospitalisation costs, and access to treatment in other countries, and especially in LMICs.

## Discussion

Dexamethasone is a globally accessible existing treatment that can be highly cost effective if given to hospitalised COVID-19 patients requiring oxygen therapy. Particularly in the UK setting, Dexamethasone is estimated to save up to 27,000 lives in the coming 6 months at a cost of up to £1640 per life year gained, which outshines the recently implemented HPV vaccination ICER^19^, e.g. In developed countries with an ageing population, where access to respiratory support is not expected to be an issue, Dexamethasone should promptly be adopted as standard treatment for patients with respiratory distress. This treatment, if given in accordance with the current guidance on patient eligibility for oxygen therapy, even where access to oxygen is limited, could save hundreds of thousands of lives in the coming six months of the pandemic. In settings where social distancing and other non-pharmaceutical interventions are becoming untenable, dexamethasone could serve both to reduce mortality and mitigate the burden on health systems.

## Data Availability

all used data is oublicly available

## Acknowledgements

We thank all members of the COVID-19 International Modelling Consortium and their collaborative partners. This work was supported by the COVID-19 Research Response Fund, managed by the Medical Sciences Division, University of Oxford. LJW is supported by the Li Ka Shing Foundation. RA acknowledges funding from the Bill and Melinda Gates Foundation (OPP1193472).

## Online Methods

We calculate the population expected to be exposed in each age class by multiplying the proportion of population expected to be exposed during the 6-month period, *α*, (**Error! Reference source not found**.) by the number of people in each age group, *n* (Supplementary Table 1). We obtain the number of people expected to require hospitalisation by multiplying the previous figure by the age structured proportion of all exposures leading to hospitalisation, *p*_*H*_, (Supplementary Table 1). We then multiply this vector by various combinations of probabilities depending on the within-hospital patient pathway (Figure 1), structured by normalised hospitalisation fatality rate, *p*_*F*_, (Supplementary Table 1) as follows:

- Expected deaths in hospitalised patients that do not require oxygen (either non-invasive or ventilator) in each age category:

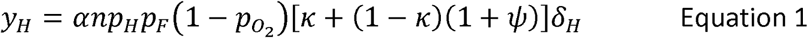
- Expected deaths in hospitalised patients who require non-invasive oxygen and receive dexamethasone in each age category:

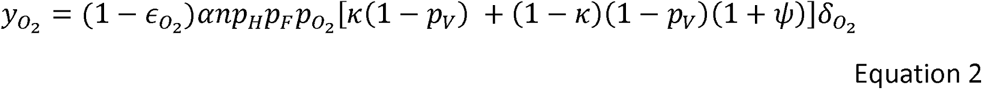
- Expected deaths in hospitalised patients who require mechanical ventilation and receive dexamethasone in each age category:

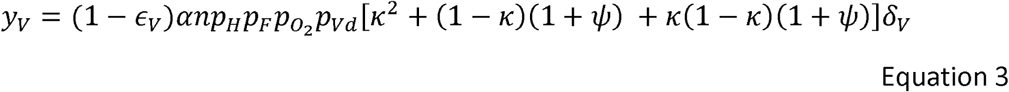

Then, the expected number of deaths in patients in each age category with severe respiratory distress that receive dexamethasone is given by:

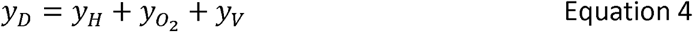

The expected number of deaths in each age category in the same patients if they do not receive dexamethasone is:

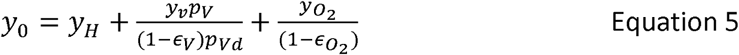

The potential lives saved, summed over all age categories, is then given by ∑ *y*_*L*_where:

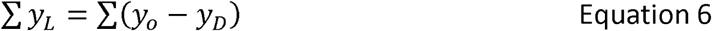

The model follows the disease pathway of COVID-19-infected patients with severe respiratory illness requiring hospitalization, non-invasive oxygen and mechanical ventilation. Depending on the strategies under investigation, patients may or may not be further split by type of management (general ward versus ICU) and use of medication (e.g. dexamethasone). In short, a proportion 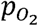 of individuals admitted to hospital with respiratory distress will require some form of oxygen. A subset of those, *p*_*Vd*_, will need mechanical ventilation. If people have perfect access to the treatment they require, their outcome is determined by a treatment specific, age-dependent probability of death. This is calculated by multiplying the age-dependent mortality modulation vector *p*_*F*_ (Supplementary Table 1) by the appropriate treatment requirement death probabilities in Table 1. If access to oxygen and/or ventilation is not perfect, the patient will not receive the most appropriate treatment, which will be reflected in an increase in mortality probability given by *ψ*. Prompt treatment with dexamethasone is assumed to reduce the need for patient ventilation by 13%, *p*_*Vd*_.

The costing analysis makes use of the simple state transition modelling framework described above (and illustrated in Figure 1). The analysis takes a provider (health system) perspective. Costs are expressed in 2020 prices, and no discounting is undertaken given that all costs within the analyses are incurred within a short period of time. The models include two absorbing states: recovered and dead. The former is assumed to have no morbidity loss (i.e. patients return to their pre-COVID-19 health state upon recovery) while the latter captures the years of life lost (YLL) for a person dying of COVID-19. The YLL associated with death is taken from secondary data and is based on estimated life expectancy for UK citizens in each of the five-year age groups^16^.

From the patient pathway model in Figure 1, we can extrapolate hospital utilisation, including the proportion of hospitalised individuals occupying ICU vs surge beds. To calculate hospitalisation costs per patient hospital pathway, we then need to know the length of stay per patient in each type of hospital bed. We assume that patients requiring oxygen alone will only occupy surge beds during their stay, regardless of outcome. However, patients that do recover spend on average 4.5 more days in hospital than those who die^15,20^,. Patients needing mechanical ventilation will occupy ICU beds for an average of 7 days if they survive and then spend an extra 5 days on average in a surge bed^15^. Ventilated patients who eventually die typically spend 8 days in an ICU bed. We then use the incremental number of days spent in hospital, together with unit cost data per inpatient day in surge beds and intensive care beds, taken from the NHS National Tariff^17^, to inform the total incremental cost of lives saved and life years gained. The costs used are provided in Supplementary Table 2. To assess the cost-effectiveness of dexamethasone treatment, we need to compute the additional cost of hospital treatment including dexamethasone as compared with the cost of treatment without dexamethasone, known as the incremental cost-effectiveness ratio (ICER). To calculate the ICER, the cost of providing dexamethasone is subtracted from the cost of treatment without dexamethasone and divided by the difference in YLL for the two treatment arms (no dexamethasone vs dexamethasone). The difference between the YLL for the two treatment arms then becomes the years of life gained (YLG).

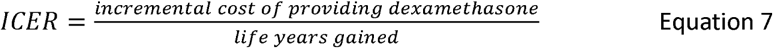

To reflect the uncertainty inherent to the estimated numbers of deaths prevented, life years gained, total incremental cost, incremental cost per life saved, and incremental cost per life year gained, we perform sensitivity analyses through Latin-hypercube sampling. Simply put, we iteratively calculate our interest outcome variables, where at each iteration we sample a value for selected input values from uniform distributions with set intervals, with ranges given in Table 1. For all sensitivity analyses we perform 1 million iterations and present the results as median and 5^th^ and 95th percentiles, i.e. the 90% double-sided confidence intervals. Throughout the manuscript, we presented epidemiological and costing estimates calculated when assuming the infection hospitalisation ratio (IHR) and the hospitalisation fatality ratio (HFR) in the UK resemble that estimated from French data^10^. Ideally, we would use UK age-dependent serology data to inform a UK specific IHR. Unfortunately, those data are not currently available so we must rely on other countries estimates/data. Whilst we believe the French estimates are a good reflection of the UK reality, we re-calculate all estimates (Supplementary Table 3), assuming HFR and HFR to follow age patterns similar to those measured in Spain (Supplementary Table 4).

**Supplementary Table 1.** Age structured values for the UK population, *n* ^9^, the infection hospitalisation rate, pH^10^, and the normalised hospitalisation fatality rate, pF^10^.

| Age group<br>(years) | Population<br>($n$) | $p_H$ | $p_F$ |
| --- | --- | --- | --- |
| 0-4 | 3,924,490 | 0.002 | 0.019 |
| 5-9 | 4,119,566 | 0.002 | 0.019 |
| 10-14 | 3,956,340 | 0.002 | 0.019 |
| 15-19 | 3,686,133 | 0.002 | 0.019 |
| 20-24 | 4,074,640 | 0.006 | 0.035 |
| 25-29 | 4,484,067 | 0.006 | 0.035 |
| 30-34 | 4,706,828 | 0.013 | 0.060 |
| 35-39 | 4,588,196 | 0.013 | 0.060 |
| 40-44 | 4,308,130 | 0.017 | 0.10 |
| 45-49 | 4,296,121 | 0.017 | 0.10 |
| 50-54 | 4,634,540 | 0.035 | 0.21 |
| 55-59 | 4,538,925 | 0.035 | 0.21 |
| 60-64 | 3,905,016 | 0.071 | 0.40 |
| 65-69 | 3,381,761 | 0.071 | 0.40 |
| 70-74 | 3,388,488 | 0.113 | 0.66 |
| 75-79 | 2,442,147 | 0.113 | 0.66 |
| 80-84 | 1,736,567 | 0.32 | 1 |
| 85-89 | 1,077,555 | 0.32 | 1 |
| 90-94 | 490,577 | 0.32 | 1 |
| 95-99 | 130,083 | 0.32 | 1 |
| 100+ | 15,834 | 0.32 | 1 |

**Supplementary Table 2:** Cost inputs and assumptions used.

| Variable | Value (GBP) |
| --- | --- |
| Dexamethasone treatment course | 5.00 |
| Cost per day for patients on mechanical ventilation in an ICU | 4,520.00 |
| Cost per day for patients on non-invasive ventilation with oxygen | 1,356.00 |
| Incremental number of days spent in hospital by survivors | 4.5 |

**Supplementary Table 3:** Sensitivity of the main epidemiological and costing metrics to assumptions on the infection hospitalisation rate, *p*_*H*_, and the normalised hospitalisation fatality rate, *p*_*F*_. The two countries from which *p*_*h*_ and *p*_*f*_ vectors are derived, and corresponding median and 90% confidence intervals are shown in columns.

|  | France | Spain |
| --- | --- | --- |
| **Lives Saved** | 12,000<br>[4,250 – 27,000] | 5,200<br>[2,000 – 12,000] |
| **Life Years Gained** | 126,000<br>[46,000 – 290,000] | 60,000<br>[20,500 – 134,000] |
| **Total Incremental Cost (£)** | 84,500,000<br>[5,700,000 – 330,000,000] | 37,500,000<br>[2,500,000 – 143,300,000] |
| **Incremental Cost per Life Saved (£)** | 8,230<br>[565 – 17, 700] | 8,200<br>[650 - 17,500] |
| **Incremental Cost per Life Year Gained (£)** | 760<br>[52 – 1,640] | 720<br>[50 - 1,550] |

**Supplementary Table 4.**
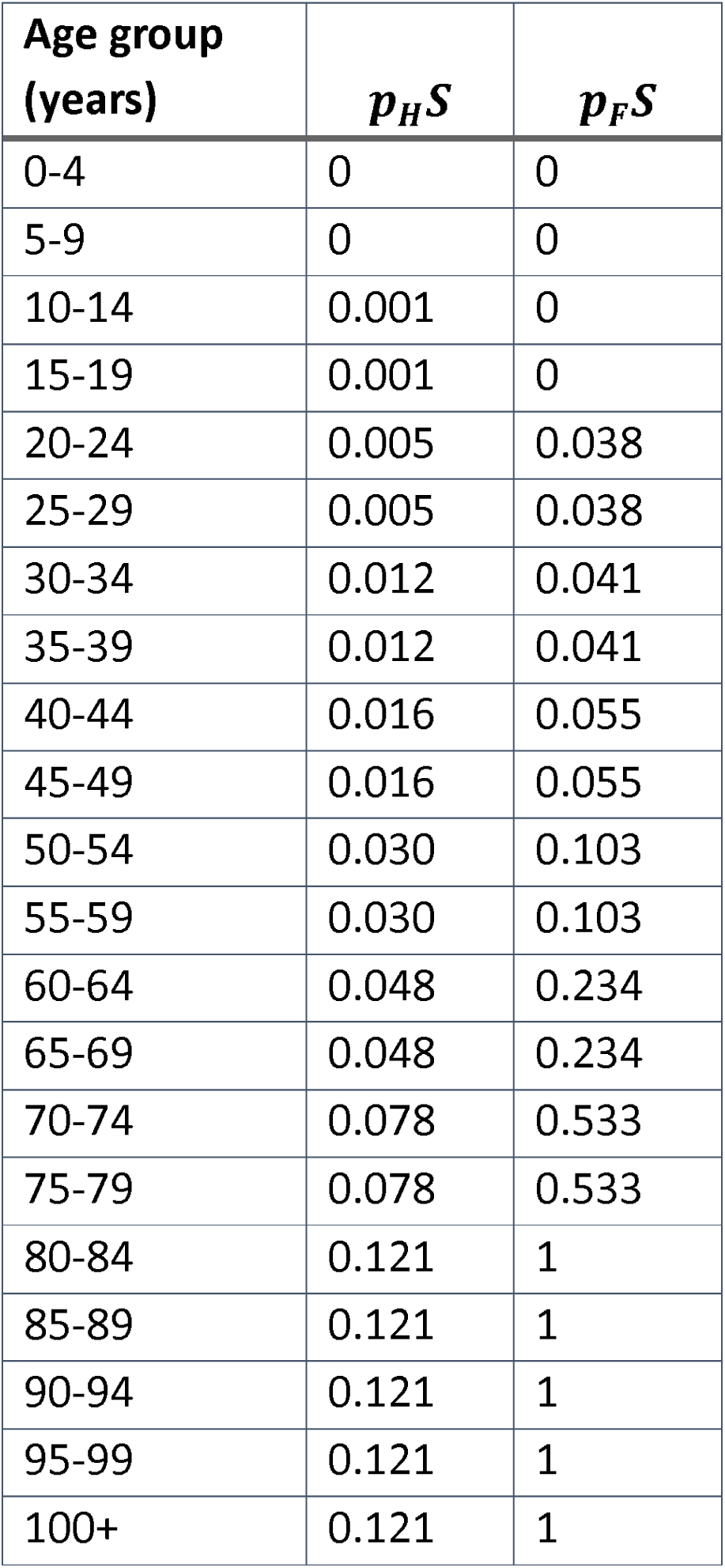
Infection hospitalisation rate, p_H_*S*, and the normalised hospitalisation fatality rate, *p*_*F*_*S* derived from Spanish data^21,22^.

| Age group<br>(years) | $p_H S$ | $p_F S$ |
| --- | --- | --- |
| 0-4 | 0 | 0 |
| 5-9 | 0 | 0 |
| 10-14 | 0.001 | 0 |
| 15-19 | 0.001 | 0 |
| 20-24 | 0.005 | 0.038 |
| 25-29 | 0.005 | 0.038 |
| 30-34 | 0.012 | 0.041 |
| 35-39 | 0.012 | 0.041 |
| 40-44 | 0.016 | 0.055 |
| 45-49 | 0.016 | 0.055 |
| 50-54 | 0.030 | 0.103 |
| 55-59 | 0.030 | 0.103 |
| 60-64 | 0.048 | 0.234 |
| 65-69 | 0.048 | 0.234 |
| 70-74 | 0.078 | 0.533 |
| 75-79 | 0.078 | 0.533 |
| 80-84 | 0.121 | 1 |
| 85-89 | 0.121 | 1 |
| 90-94 | 0.121 | 1 |
| 95-99 | 0.121 | 1 |
| 100+ | 0.121 | 1 |

## CoMo Consortium authors

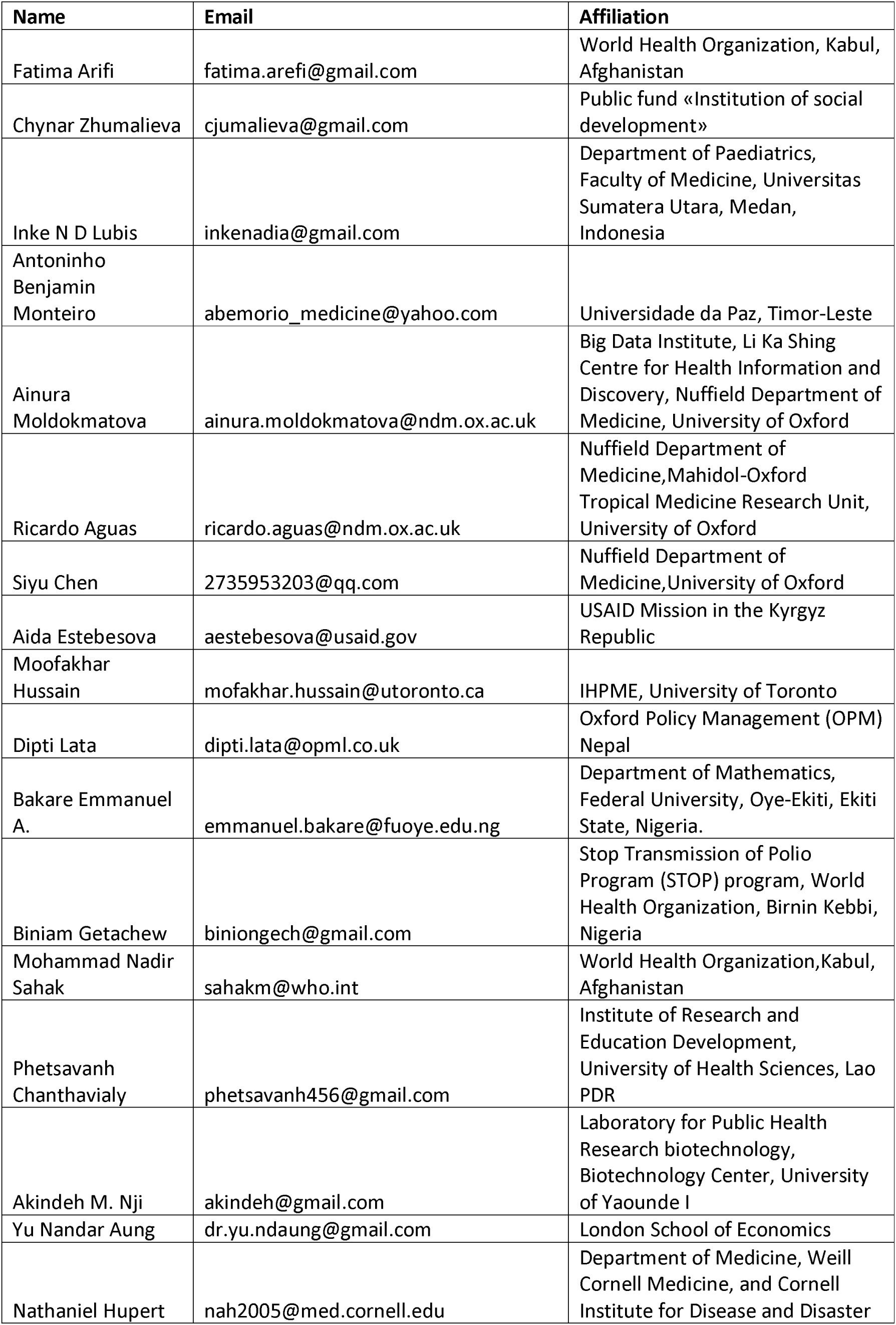
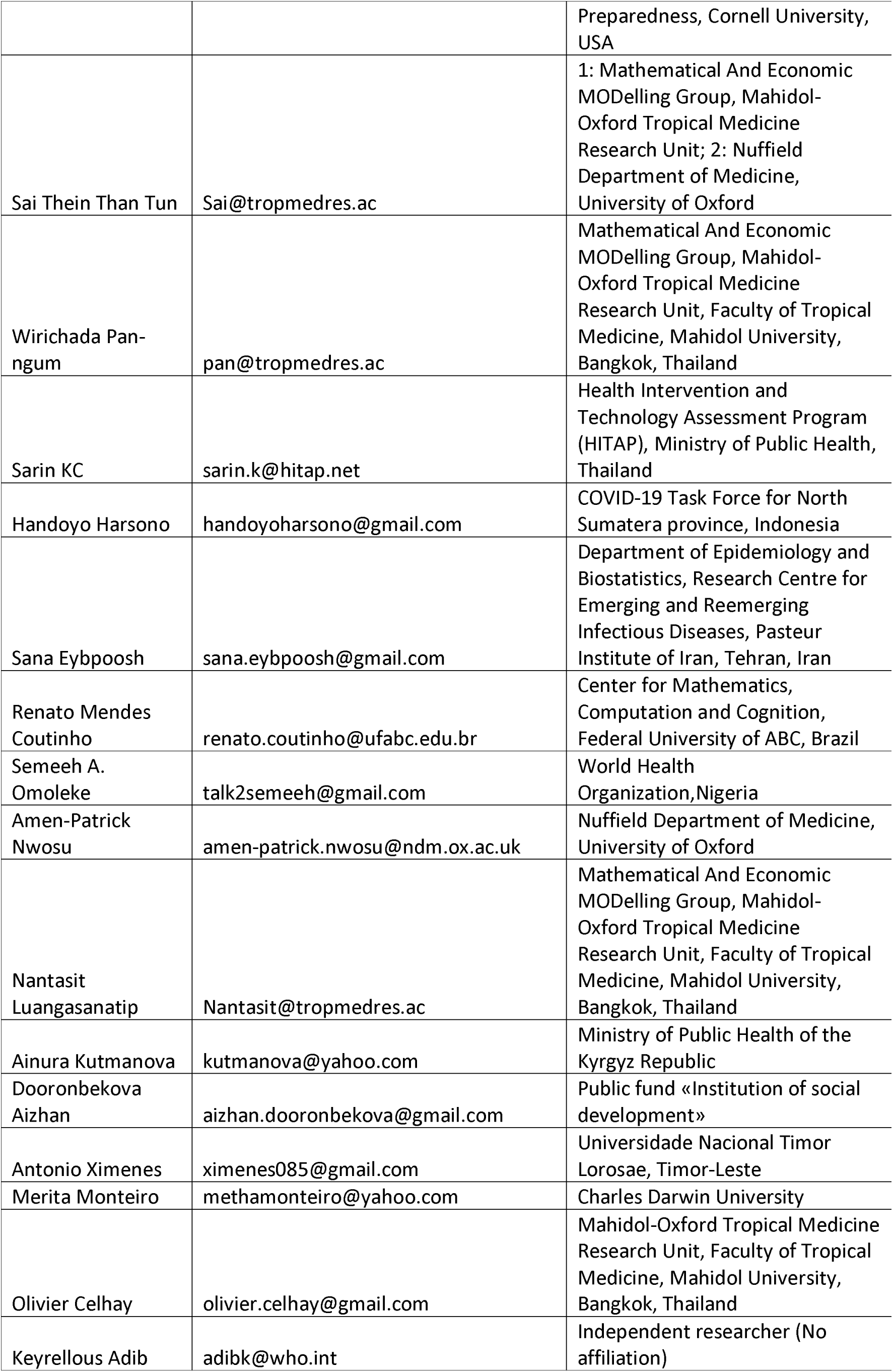
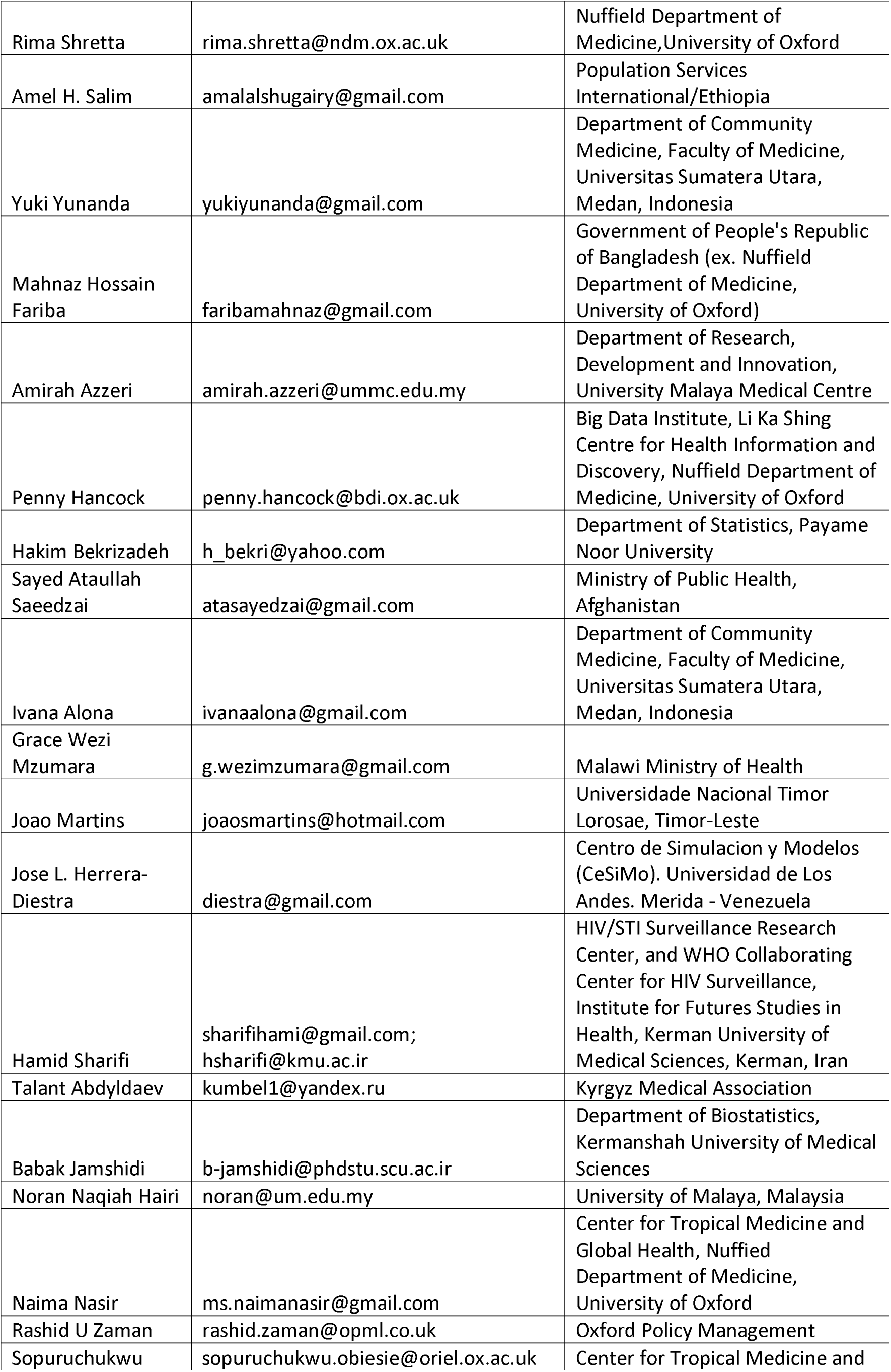
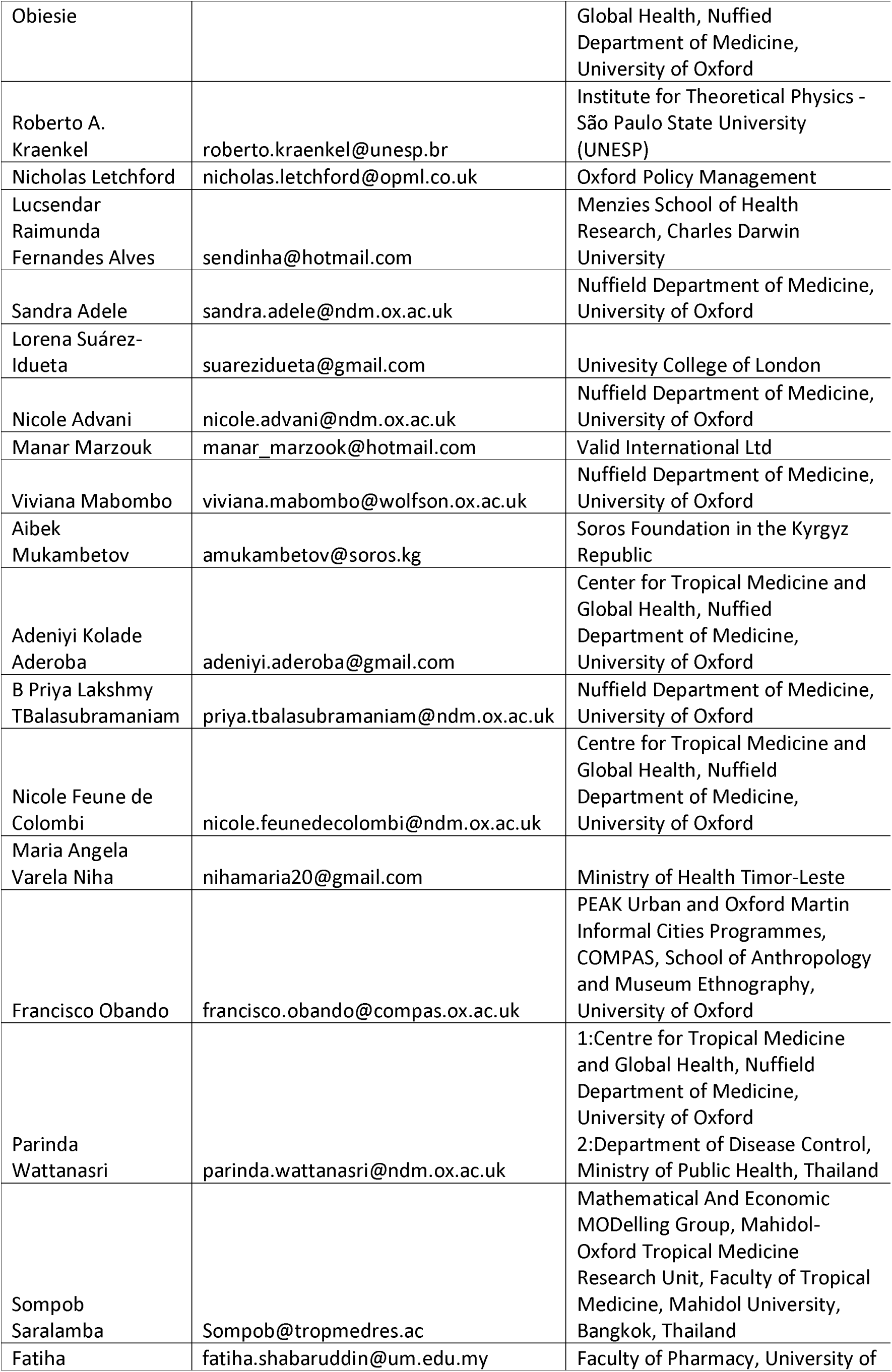
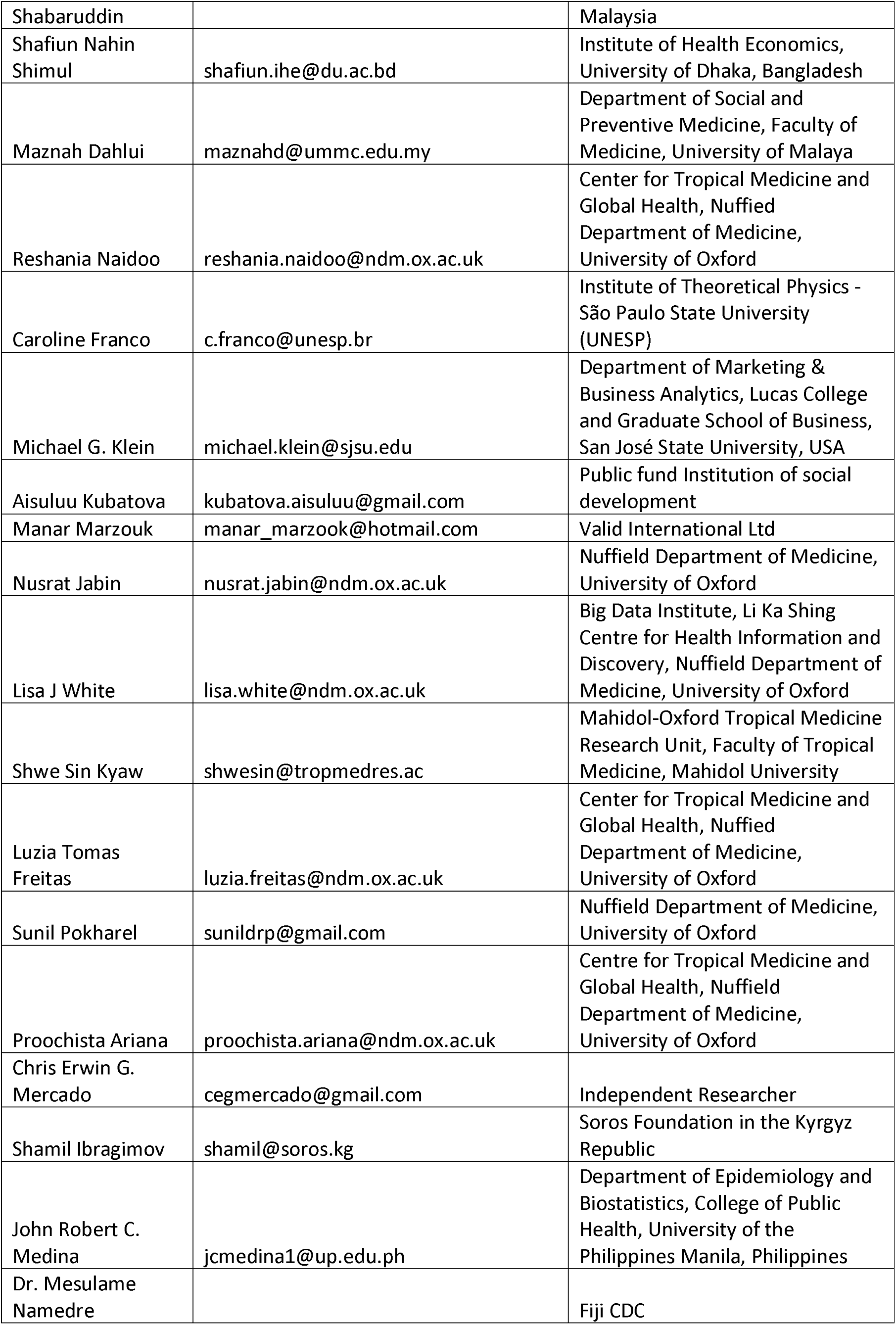

